# Prospective evaluation of dRAST for selecting optimal targeted antibiotics in positive blood culture with Gram-positive organisms

**DOI:** 10.1101/2022.11.06.22282012

**Authors:** Jeong-Han Kim, Taek Soo Kim, Chang Kyung Kang, Sangkwon Han, Dong Young Kim, Sunghoon Kwon, Pyeong Gyun Choe, Nam Joong Kim, Wan Beom Park, Myoung-don Oh

## Abstract

Empirical antibiotic selection often fails to be optimal targeted in the era of increasingly common resistant organisms. We prospectively evaluated the usefulness or rapid AST for optimal antibiotic selection by infectious disease (ID) physicians in patients with bacteremia of Gram-positive organisms. QMAC-dRAST results led to optimal antibiotic treatment in 33 (89.2%) of the 37 cases receiving non-optimal targeted antibiotics. Optimal targeted treatments based on QMAC-dRAST results were possible in 133 (97.1%) of the 137 cases. In conclusion, the introduction of rapid phenotypic AST can help increase the selection of optimal targeted antibiotics during the early period of bacteremia.

## Introduction

Sepsis can cause morbidity, mortality, prolonged hospitalization and high costs for healthcare systems and the most important life-saving action is the prompt administration of an appropriate antibiotic agent.^1-5^ Clinicians need the definitive identification of the pathogen and antimicrobial susceptibility testing (AST) to adjust antibiotics more precisely and effectively. Since the conventional method for identification and AST takes at least 2 – 3 days, they start an empirical treatment at the beginning of blood stream infection (BSI) diagnosis and adjust antibiotics based on Gram stain results which is still empirical.^6-7^ However, those empirical treatments often fail to guide for selecting optimal targeted antibiotics in the era of increasingly common resistant organisms.

To provide ID and AST results at the early stage of BSI, many rapid diagnostic tests have been developed directly using positive blood cultures. MALDI-TOF MS has been established for rapid pathogen identification and automated molecular diagnostic tests can identify both pathogens and resistance genes.^8-11^ Moreover, several rapid phenotypic AST systems have been developed.^12-14^ Especially, dRAST is a phenotypic AST system for Gram-positive cocci as well as Gram-negative rods utilizing microfluidic technology for bacteria immobilization and microscopic image analysis of bacteria growth.^14^

In a recent study, we evaluated how well MALDI-TOF MS and dRAST guide ID physicians in the selection of optimal targeted antibiotics for patients with positive blood cultures.^15^ Here, more specifically, we showed the results for blood stream infection with Gram-positive organisms.

## Methods

The current analysis was performed using the data presented in the previous clinical study.^15^ The data were separated into Gram-positive and Gram-negative cases and then the antibiotic treatments decided by ID physicians based on each BSI diagnosis result were categorized as follows: optimal targeted, unnecessary broad spectrum, suboptimal and ineffective as previously defined.^15^ To compare with more early stage empirical treatment, we added empirical antibiotics treated at the time of blood collection for blood cultures in this analysis.

## Impact of dRAST on ID physician antibiotic selection

In initial empirical antibiotic selection, the ID physicians chose optimal targeted antibiotics in 26 (19.0%) cases, unnecessary broad-spectrum antibiotics in 27 (19.7%) cases and suboptimal or ineffective antibiotics 84 (61.3%) cases (Table 1). After reviewing the Gram stain results, the ID physicians chose optimal targeted antibiotics in 83 (60.6%) cases, unnecessary broad-spectrum antibiotics in 20 (14.6%) cases and suboptimal or ineffective antibiotics in 34 (24.8%) case. After reviewing MALDI-TOF MS results, the ID physicians chose optimal targeted antibiotics in 100 (73.0%) cases, unnecessary broad-spectrum antibiotics in 11 (8.0%) cases and ineffective antibiotics in 26 (19.0%) cases. The MALDI-TOF MS results enabled appropriate antibiotic selection in 119 (86.9%) cases, but the proportion of ineffective antibiotic selection was high for resistant strains (23.1%) and the proportion of unnecessary broad-spectrum antibiotic selection was high for susceptible strains (18.6%).

After reviewing the dRAST results, the ID physicians chose optimal antibiotic treatment for 33 (89.2%) of the 37 patients who were recommended non-optimal antibiotics based on the MALDI-TOF MS results. Specifically, there was no ineffective antibiotic selection for resistant strains and unnecessary broad-spectrum antibiotic selection for susceptible strains was reduced to 2 (3.4%) cases. Overall, optimal targeted treatments based on the dRAST results were possible in 133 (97.1%) of the 137 cases and the proportion of it was not significantly different between susceptible strains and resistant strains.

With the steady increase in resistant strains such as methicillin-resistant *Staphylococcus aureus* (MRSA) and vancomycin-resistant *Enterococcus* species (VRE), an empirical antibiotic selection is highly possible to be ineffective. And as shown in our data, the rapid pathogen identification alone didn’t play well a role of guiding the empirical antibiotic treatment for resistant strains. In this regards, rapid phenotypic AST should be introduced and used for antibiotic decision-making.

Rapid molecular diagnostic methods enable the rapid detection of antibiotic resistance genes of Gram-positive organisms and may greatly assist with antibiotic selection.^10,11,16,17^ However, because they mostly detect mecA/C and vanA/B only and can’t tell the susceptibility of the other antibiotics, it is hard to decide de-escalation or escalation of antibiotics properly. In contrast, rapid phenotypic AST gives susceptibility results of various antibiotics and this provides better clinical usefulness.

## Conclusion

Our present data show that not only empirical antibiotic selection but also antibiotic selection based on rapid pathogen identification may frequently be inadequate or unnecessary broad-spectrum for Gram-positive organisms. With the increasing number of resistant organisms in clinical practice, the introduction of rapid phenotypic AST can help increase the selection of optimal targeted antibiotics during the early period of bacteremia.

**Figure 1.**
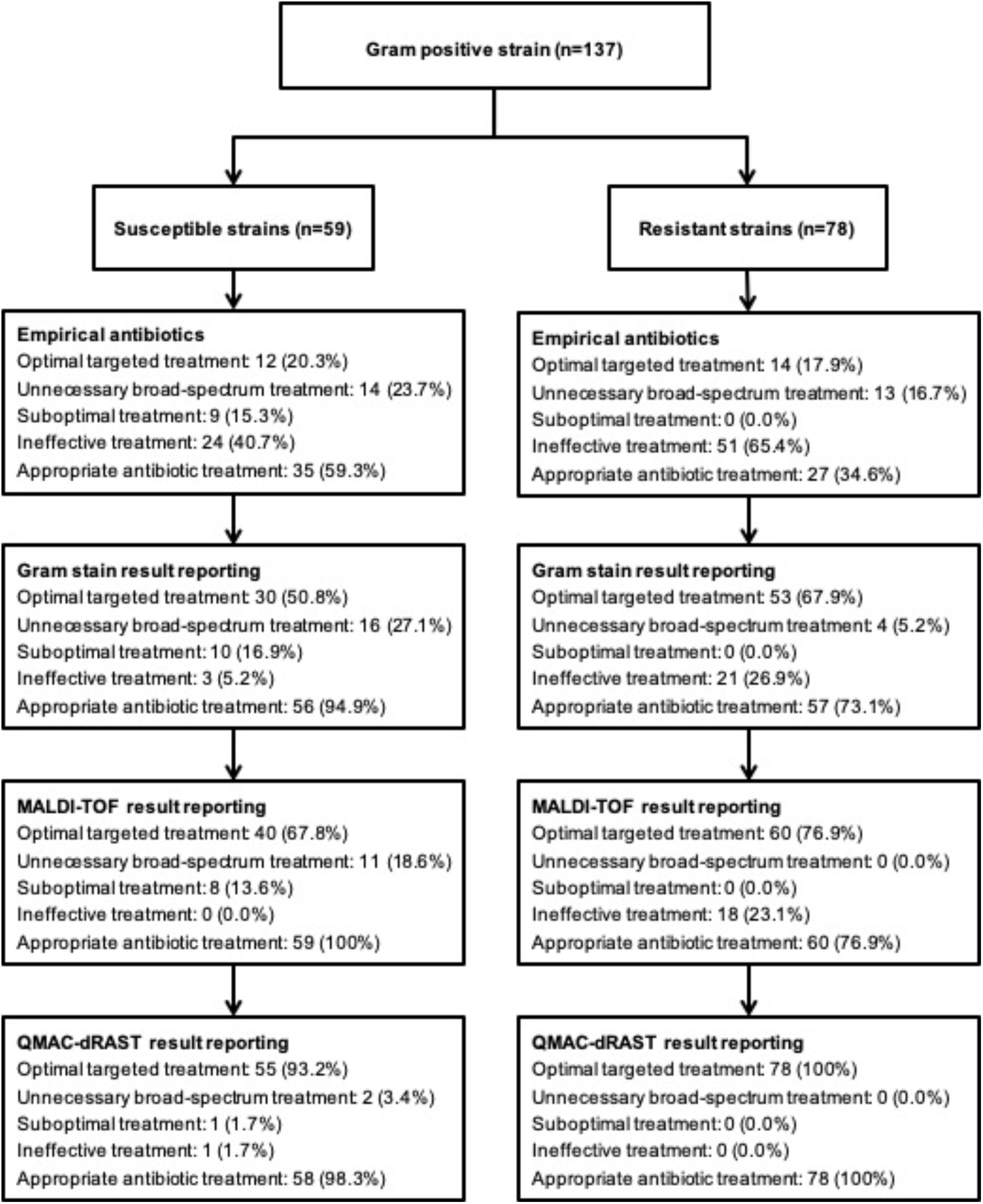
ID physician antibiotic selection based on the results of Gram staining, MALDI-TOF MS and dRAST methods. Resistant strains include MRSA, VRE and MDR strains. Optimal targeted treatment: the most effective and narrowest spectrum antibiotic treatment for the pathogen(s). Unnecessary broad-spectrum treatment: antibiotic treatment that is effective against the pathogen(s), but has broad-spectrum activity requiring de-escalation. Suboptimal treatment: antibiotic treatment to which the pathogen(s) are susceptible, but that has inferior antimicrobial activity compared with that of the optimal targeted treatment. Ineffective treatment: antibiotic treatment to which the pathogen(s) have intermediate susceptibility or are resistant. Appropriate antibiotic treatment: antibiotic treatment to which the pathogen(s) are susceptible.

## Data Availability

All data produced in the present study are available upon reasonable request to the authors

## References

1. Pittet D, Tarara D, Wenzel RP. Nosocomial blood stream infection in critically ill patients. Excess length of stay, extra costs, and attributable mortality. JAMA 1994; 271: 1598–601.

2. Arefian H, Heublein S, Scherag A et al. Hospital-related cost of sepsis: a systematic review. J Infect 2017; 74: 107–17.

3. Ibrahim EH, Sherman G, Ward S et al. The influence of inadequate antimicrobial treatment of bloodstream infections on patient outcomes in the ICU setting. Chest 2000; 118: 146–55.

4. Kumar A, Roberts D, Wood KE et al. Duration of hypotension before initiation of effective antimicrobial therapy is the critical determinant of survival in human septic shock. Crit Care Med 2006; 34: 1589–96.

5. Rhodes A, Evans LE, Alhazzani W et al. Surviving sepsis campaign: international guidelines for management of sepsis and septic shock: 2016. Intensive Care Med 2017; 43: 304–77.

6. Choi J, Yoo J, Lee M et al. A rapid antimicrobial susceptibility test based on single-cell morphological analysis. Sci TranslMed 2014; 6: 267ra174.

7. Munson EL, Diekema DJ, Beekmann SE et al. Detection and treatment of blood stream infection: laboratory reporting and antimicrobial management. J Clin Microbiol 2003; 41: 495–7.

8. Seng P, Drancourt M, Gouriet F et al. Ongoing revolution in bacteriology: routine identification of bacteria by matrix-assisted laser desorption ionization time-of-flight mass spectrometry. Clin Infect Dis 2009; 49: 543–51.

9. Huang AM, Newton D, Kunapuli A et al. Impact of rapid organism identification via matrix-assisted laser desorption/ionization time-of-flight combined with antimicrobial stewardship team intervention in adult patients with bacteremia and candidemia. Clin Infect Dis 2013; 57: 1237–45.

10. Bork JT, Leekha S, Heil EL et al. Rapid testing using the Verigene Gram negative blood culture nucleic acid test in combination with antimicrobial stewardship intervention against Gram-negative bacteremia. Antimicrob Agents Chemother 2015; 59: 1588–95.

11. Salimnia H, Fairfax MR, Lephart PR et al. Evaluation of the FilmArray blood culture identification panel: results of a multicenter controlled trial. J Clin Microbiol 2016; 54: 687–98.

12. Charnot-Katsikas A, Tesic V, Love N et al. Use of the Accelerate Pheno System for identification and antimicrobial susceptibility testing of pathogens in positive blood cultures and impact on time to results and workflow. J Clin Microbiol 2017; 56: e01166–17.

13. Marschal M, Bachmaier J, Autenrieth I et al. Evaluation of the Accelerate Pheno System for fast identification and antimicrobial susceptibility testing from positive blood cultures in blood stream infections caused by Gram-negative pathogens. J Clin Microbiol 2017; 55: 2116–26.

14. Choi J, Jeong HY, Lee GY et al. Direct, rapid antimicrobial susceptibility test from positive blood cultures based on microscopic imaging analysis. Sci Rep 2017; 7: 1148.

15. Kim J-H, Kim TS, Jung H et al. Prospective evaluation of a rapid antimicrobial susceptibility test (QMAC-dRAST) for selecting optimal targeted antibiotics in positive blood culture. J Antimicrob Chemother 2019; 74: 2255–2260

16. Kaleta EJ, Clark AE, Johnson DR et al. Use of PCR coupled with electrospray ionization mass spectrometry for rapid identification of bacterial and yeast bloodstream pathogens from blood culture bottles. J Clin Microbiol 2011; 49: 345–53.

17. Evans SR, Hujer AM, Jiang H et al. Rapid molecular diagnostics, antibiotic treatment decisions, and developing approaches to inform empiric therapy: PRIMERS I and II. Clin Infect Dis 2016; 62: 181–9.

